# Human papillomavirus intermittence, redetections, and associated risk of cytological abnormalities in the Ludwig-McGill cohort study of adult women

**DOI:** 10.1101/2022.10.12.22280699

**Authors:** Talía Malagón, Helen Trottier, Mariam El-Zein, Luisa L Villa, Eduardo Franco, the Ludwig-McGill Cohort Study

## Abstract

**Introduction:** We assessed the incidence of redetection with the same human papillomavirus (HPV) genotype, predictors of first HPV detections and redetections, and prevalence of cytological lesions during HPV redetections.

**Methods:** The Ludwig-McGill cohort study followed women aged 18-60 years from São Paulo, Brazil in 1993-1997 for up to 10 years. Women provided cervical samples for cytology testing and HPV DNA testing at each visit. A redetection was defined as a recurring genotype-specific HPV positive result after one or more intervening negative visits. Predictors of genotype-specific redetection were assessed using adjusted hazard ratios (aHR) with Cox regression modeling.

**Results:** 2184 women contributed 2368 incident HPV genotype-specific first detections and 308 genotype-specific redetections over a median follow-up of 6.5 years. The cumulative incidence of redetection with the same genotype was 7% at 1 year and 15% at 5 years after the loss of positivity of the first detection. Neither age (aHR 0.90, 95%CI 0.54-1.47 for ≥45y vs. <25y) nor new sexual partner acquisition (aHR 0.98, 95%CI 0.70-1.35) were statistically associated with genotype-specific redetection. High-grade squamous intraepithelial lesion prevalence was similar during first HPV detections (2.9%) and redetection (3.2%).

**Conclusions:** Our findings suggest many HPV redetections were likely reactivations of latent recurring infections.

## Introduction

Human papillomavirus (HPV) DNA testing is the primary screening test for cervical cancer in many countries, and is the test recommended by the World Health Organisation for cervical cancer screening worldwide.[1] As increasingly many women worldwide are testing positive for HPV through screening, it is more crucial than ever to understand the course of HPV infection over a woman’s lifetime. The emerging model of HPV natural history holds that while initial genital HPV infection is generally acquired via sexual exposure in adolescence or young adulthood, the course of infection after initial acquisition can be non-linear and follow different pathways.[2] The loss of HPV DNA detectability in cervical samples may not always reflect true immune clearance. In some cases, it may occur due to immune control of the infection below the limit of detection in a state of viral latency.[3] Consequently, the redetection of HPV DNA after a period of negativity could be due to several reasons, including true new reinfection from a sexual exposure, intermittent detection of a latent infection, or simply transient deposition from a cross-infection at another epithelial site or from a recent sex act.

Many questions remain regarding HPV redetections after periods of test negativity, including what is the long-term risk of redetection, whether the risk of HPV redetection changes with age, and whether a redetection of HPV confers the same risk of cervical cancer as the initial infection detection.[2] Most natural history studies have had insufficient sample sizes and follow-up to study redetections of HPV. These questions are of increasing clinical importance, as many women may present with recurring HPV positive results throughout their lives, and it is likely that in the future the screening management of a woman with HPV-positive results may depend on her previous history of HPV positivity.[4-6]

The Ludwig-McGill cohort study was a longitudinal study of the natural history of HPV infection and cervical neoplasia which has contributed much to our knowledge of HPV natural history over the past thirty years.[7-11] We have previously published an analysis of the HPV redetections that were most likely to be true new reinfections from new sexual exposures; that analysis was restricted to redetections which occurred after three intervening negative test results.[8] However, these redetections only represented a small fraction of all redetections that occurred in the cohort. Our objective with this new analysis was to examine all redetections of the same HPV genotype in women to assess how frequently HPV genotype-specific redetections occur, what are the predictors of HPV redetection in women who have previously tested HPV positive, and whether redetections are associated with the same risk of cytological lesions as first HPV detections.

## Methods

### Study design and participants

The study protocol and methods have been previously described in detail elsewhere.[7] Briefly, the study recruited women attending a maternal and child health program catering to low-income families in the city of São Paulo, Brazil between 1993-1997. Women were eligible to participate if they were between 18 and 60 years old, were not currently pregnant and had no intention of becoming pregnant during the next 12 months, had an intact uterus, reported no use of vaginal medication in the previous 2 days, and had not had treatment for cervical disease in the previous 6 months.

Women were followed-up every 4 months during the first year, and subsequently twice a year for up to 10 years after enrolment. During the first four visits and at every second visit thereafter, women completed an interviewer-administered questionnaire on demographic, socioeconomic, and behavioral risk factors for HPV infection and cervical cancer. Study personnel collected cervical samples for cytology and HPV DNA testing at each visit. Meal tickets were provided to the women as incentives to remain in the study.

All women provided signed consent forms, and ethical approval was obtained from the institutional review boards of McGill University (Montreal, Canada), University of Toronto (Toronto, Canada), Ludwig Institute for Cancer Research (São Paulo, Brazil), and Maternidade Escola Dr. Mario de Moraes Altenfelder Silva Municipal Hospital clinic (São Paulo, Brazil).

### Cervical smears

Ectocervical and endocervical cells were collected using an Accelon biosampler (Medscand Inc., Hollywood, FL, USA) and placed in a tube containing Tris-EDTA buffer. Cervical smears were prepared on a glass slide and fixed in 95% ethanol, stained, and read at the São Paulo Branch of the Ludwig Institute’s cytopathology laboratory for initial diagnosis based on the Papanicolaou system. The smears were then shipped to Montreal and re-read at the Jewish General Hospital based on the Bethesda system.[12] Women who had moderate or worse dysplasia based on initial readings with the Papanicolaou system or who had high-grade squamous intraepithelial lesions (HSIL) on subsequent readings with the Bethesda system were referred for colposcopy and management according to the local prevailing protocol. For this analysis, we used the Montreal cytology results based on the Bethesda system.

### HPV genotyping

HPV DNA was extracted, purified by spin column chromatography, and amplified by PCR, using the MY09/11 and PGMY protocols.[13-15] Typing of the amplified products was performed by hybridization with individual oligonucleotide probes for 27 HPV genotypes, and restriction fragment length polymorphism for the samples that hybridized with the generic but none of the genotype-specific probes.[16] These combined methods allowed the identification of over 40 genital HPV genotypes, including unknown genotypes. We classified as high-risk oncogenic HPV genotypes 16, 18, 31, 33, 35, 39, 45, 51, 52, 56, 58, 59, 66, and 68, based on both carcinogenicity[17] and for comparability as these genotypes are the targets of many commercial HPV tests. In order to check the integrity of samples, the assays also included an additional set of primers (GH20 and PC04) to amplify a 268 bp region of the β-globin gene.[13] Samples that were negative for both HPV and β-globin were considered invalid and excluded from analyses.

### Statistical analysis

All analyses were genotype-specific, with the unit of observation being the HPV genotype. Hence, women contributed multiple observations to analyses corresponding to individual HPV genotypes. Results from all individual HPV genotypes were pooled together, except where results are presented by genotype.

To analyze detection patterns, we examined women with at least 3 study visits. We considered HPV genotypes to be prevalent if they were detected at the study baseline, and incident if detected at subsequent visits in women who were negative at baseline for that HPV genotype. We considered a genotype-specific HPV detection pattern to be persistent when the same genotype was detected at two or more successive visits with valid HPV test results, transient when a genotype was detected only at a single visit, and intermittent when a genotype was detected at two or more visits separated by at least one intervening negative test for that genotype.

To calculate cumulative HPV detection incidence, we analyzed women with at least 2 study visits using stratified Cox proportional hazards models for repeated events. The models were stratified, with different baseline hazard functions for the first, second, and third or more detections with the same HPV genotype. The time to first genotype-specific HPV detection was modeled from the baseline study visit in women who were genotype-specific negative at baseline. The time to genotype-specific HPV redetection (second or more detections) was modeled from the date that a woman became negative for a genotype she was previously positive for (the “clearance” date). We used the robust sandwich estimate of Lin and Wei to account for having multiple observations from the same woman.[18] The cumulative incidence of first genotype-specific detection and re-detection overall and by age were estimated from the survivor functions of stratified Cox models.

For regression models, we considered as *a priori* predictors demographic and socioeconomic variables (age, menopausal status, income quartile, race) and behavioral variables (smoking, number of lifetime sex partners, new sex partners, current sex partners, condom use) known to be associated with HPV acquisition. Age and behavioral variables were analyzed as time-varying exposures. We modeled each variable individually in a univariate model, and all together in a multivariable model. Models had interaction terms to allow predictors to have different effects for first incident detections and for redetections. We used the joint tests for the interaction terms to assess whether hazard ratios (HR) differed for first incident detections compared with redetections, and whether rates of first detection and redetection varied by HPV genotype.

Values for some predictors were missing at some visits either due to non-response or design (questionnaires were given only every second visit after the first year, and questions on income, race, and menopausal status were not repeated in later questionnaires). For sexual behavior variables, we imputed the woman’s last non-missing response backwards to previous visits with missing data. For other variables, we imputed the woman’s last non-missing response forward to subsequent visits with missing data.

Finally, we assessed in all women whether the prevalence of low grade squamous intraepithelial lesions or worse (LSIL), HSIL, and results of atypical squamous cells of undetermined significance or worse (ASCUS+, which includes LSIL and HSIL), differed between visits with first HPV genotype detections and redetections for high-risk HPV genotypes.

## Results

There were 2462 eligible women recruited in the Ludwig-McGill cohort study, of which 2184 (89%) had at least 2 study visits and 1986 (81%) had at least 3 study visits with valid HPV DNA data. The mean age at baseline was 32.7 years (standard deviation 8.8) and the majority of women were either married (48%) or living as married (34%). The median follow-up time for women with at least 2 study visits was 6.5 years (1^st^-3^rd^ quartile 4.3-7.8 years). In women with at least 3 study visits, 15% (60/402) of prevalent genotype-specific detections had an intermittent detection pattern, and 8% (196/2346) of incident genotype-specific detections had an intermittent detection pattern, where the same genotype was re-detected at a later visit after a negative result (Table 1).

**Table 1.** HPV genotype-specific detection patterns in women with 3 or more study visits, pooled over all HPV genotypes.

| Detection pattern | Prevalent detections<br>(N=402) |  | Incident detections<br>(N=2346) |  | Total (N=2748) |  |
| --- | --- | --- | --- | --- | --- | --- |
|  | Pattern<sup>a</sup> | n (%) | Pattern<sup>a</sup> | n (%) |  |  |
| Persistent | 111/110 | 137 (34%) | 011 | 494 (21%) | 631 | (23%) |
| Transient | 100 | 205 (51%) | 010/001 | 1656 (71%) | 1861 | (68%) |
| Intermittent | 101 | 60 (15%) | 0101 | 196 (8%) | 256 | (9%) |

<sup>a</sup>Short list of illustrative patterns (1=HPV-positive, 0=HPV-negative) over 3-4 visits that fit into each detection pattern definition. A full list of patterns is provided in Supplementary Table 1.

There were 256 second detections and 52 third or more detections of the same genotype contributing to analyses of cumulative incidence of HPV genotype-specific detections, for a total of 308 genotype-specific redetections (Figure 1). The cumulative incidence of first detection with an HPV genotype in women negative for that HPV genotype at baseline was 1% (95%CI 1-1%) 1 year after baseline, and 2% (95%CI 2-2%) 5 years after baseline, when pooled across all HPV genotypes (Figure 1, Table 2). The cumulative incidence of the second detection (first redetection) of the same genotype was 7% (95%CI 6-8%) 1 year after the date of loss of positivity of the first detection, and 15% (95%CI 13-17%) 5 years after the date of loss of positivity of the first detection, when pooled across all HPV genotypes. The cumulative incidence of additional third or more detections (all subsequent redetections) of the same genotype was 16% (95%CI 10-21%) 1 year after the date of loss of positivity of the previous detection, and 42% (95%CI 30-52%) 5 years after the loss of positivity date of the previous detection, when pooled across all HPV genotypes.

**Table 2.** Cumulative incidence of genotype-specific first HPV detection from baseline, and of same genotype redetection after at least one negative visit following the first detection in women with 2 or more study visits.

| HPV genotype | Events |  | 1-year cumulative incidence |  |  |  | 5-year cumulative incidence |  |  |  |
| --- | --- | --- | --- | --- | --- | --- | --- | --- | --- | --- |
|  | First detection<sup>a</sup> | Second detection<sup>b</sup> | First detection<sup>a</sup> |  | Second detection<sup>b</sup> |  | First detection<sup>a</sup> |  | Second detection<sup>b</sup> |  |
|  | n | n | % | (95% CI) | % | (95% CI) | % | (95% CI) | % | (95% CI) |
| All genotypes<sup>c</sup> | 2368 | 256 | 1% | (1-1%) | 7% | (6-8%) | 2% | (2-2%) | 15% | (13-17%) |
| High-risk genotypes<sup>c,d</sup> | 1145 | 116 | 1% | (1-1%) | 6% | (4-7%) | 3% | (3-4%) | 14% | (12-17%) |
| HPV6/11 | 81 | 5 | 1% | (0-1%) | 3% | (0-6%) | 3% | (2-4%) | 7% | (0-14%) |
| HPV16 | 236 | 35 | 3% | (2-4%) | 8% | (4-11%) | 10% | (8-11%) | 19% | (13-25%) |
| HPV18 | 69 | 7 | 1% | (0-1%) | 4% | (0-9%) | 3% | (2-3%) | 12% | (3-19%) |
| HPV26 | 17 | 2 | 0% | (0-0%) | 0% | (0-0%) | 1% | (0-1%) | 16% | (0-35%) |
| HPV31 | 77 | 8 | 1% | (1-2%) | 7% | (1-13%) | 3% | (2-4%) | 11% | (3-19%) |
| HPV32 | 9 | 0 | 0% | (0-0%) | 0% | NA | 0% | (0-1%) | 0% | NA |
| HPV33 | 44 | 1 | 1% | (0-1%) | 3% | (0-7%) | 2% | (1-2%) | 3% | (0-7%) |
| HPV34 | 2 | 0 | 0% | (0-0%) | 0% | NA | 0% | (0-0%) | 0% | NA |
| HPV35 | 82 | 7 | 1% | (0-1%) | 7% | (1-14%) | 3% | (2-4%) | 15% | (1-28%) |
| HPV39 | 46 | 3 | 0% | (0-0%) | 3% | (0-9%) | 1% | (1-2%) | 14% | (0-27%) |
| HPV40 | 38 | 7 | 0% | (0-1%) | 11% | (0-20%) | 1% | (1-2%) | 23% | (6-37%) |
| HPV42 | 32 | 3 | 0% | (0-0%) | 10% | (0-23%) | 1% | (1-1%) | 24% | (0-47%) |
| HPV44 | 90 | 11 | 1% | (0-1%) | 9% | (2-15%) | 3% | (2-4%) | 22% | (6-36%) |
| HPV45 | 78 | 5 | 0% | (0-1%) | 5% | (0-11%) | 3% | (2-3%) | 11% | (1-20%) |
| HPV51 | 135 | 12 | 1% | (1-2%) | 5% | (1-9%) | 6% | (5-7%) | 12% | (5-19%) |
| HPV52 | 100 | 12 | 1% | (1-1%) | 4% | (0-9%) | 3% | (2-4%) | 17% | (7-26%) |
| HPV53 | 146 | 23 | 2% | (2-3%) | 7% | (3-11%) | 6% | (4-7%) | 18% | (10-25%) |
| HPV54 | 70 | 8 | 1% | (0-1%) | 7% | (0-13%) | 3% | (2-4%) | 15% | (5-25%) |
| HPV56 | 60 | 4 | 1% | (0-1%) | 6% | (0-12%) | 2% | (2-3%) | 8% | (0-15%) |
| HPV57 | 6 | 0 | 0% | (0-0%) | 0% | NA | 0% | (0-0%) | 0% | NA |
| HPV58 | 98 | 13 | 1% | (1-1%) | 5% | (0-9%) | 4% | (3-5%) | 22% | (10-32%) |
| HPV59 | 60 | 5 | 1% | (0-1%) | 6% | (0-12%) | 2% | (1-3%) | 11% | (1-20%) |
| HPV61 | 49 | 6 | 1% | (0-1%) | 8% | (0-16%) | 2% | (1-2%) | 13% | (3-22%) |
| HPV62 | 60 | 12 | 0% | (0-1%) | 16% | (4-26%) | 2% | (2-3%) | 33% | (14-48%) |
| HPV66 | 54 | 3 | 0% | (0-1%) | 4% | (0-10%) | 2% | (1-2%) | 7% | (0-14%) |
| HPV67 | 9 | 0 | 0% | (0-1%) | 0% | NA | 0% | (0-1%) | 0% | NA |
| HPV68 | 60 | 4 | 1% | (0-1%) | 5% | (0-11%) | 2% | (2-3%) | 8% | (0-16%) |
| HPV69 | 6 | 0 | 0% | (0-0%) | 0% | NA | 0% | (0-0%) | 0% | NA |
| HPV70 | 36 | 4 | 0% | (0-1%) | 7% | (0-15%) | 1% | (1-2%) | 10% | (0-20%) |
| HPV71 | 38 | 4 | 0% | (0-1%) | 7% | (0-17%) | 1% | (0-1%) | 20% | (0-37%) |
| HPV72 | 11 | 4 | 0% | (0-0%) | 25% | (0-46%) | 0% | (0-1%) | 36% | (0-60%) |
| HPV73 | 54 | 1 | 0% | (0-1%) | 0% | (0-0%) | 2% | (1-3%) | 2% | (0-7%) |
| HPV81 | 29 | 6 | 0% | (0-1%) | 17% | (2-30%) | 1% | (1-1%) | 23% | (4-38%) |
| HPV82 | 24 | 0 | 0% | (0-1%) | 0% | NA | 1% | (0-1%) | 0% | NA |
| HPV83 | 53 | 1 | 0% | (0-1%) | 3% | (0-8%) | 2% | (1-2%) | 3% | (0-8%) |
| HPV84 | 112 | 16 | 1% | (0-1%) | 10% | (3-16%) | 4% | (3-4%) | 20% | (10-29%) |
| HPV89 | 24 | 2 | 0% | (0-0%) | 9% | (0-21%) | 1% | (0-1%) | 9% | (0-21%) |

CI=confidence interval; HPV=human papillomavirus; NA=not available (could not be calculated).
<sup>a</sup> First incident detection of a given HPV genotype from the baseline study visit.
<sup>b</sup> Second detection of the same HPV genotype from the first negative visit following first positivity with that genotype.
<sup>c</sup> Results from all individual HPV genotypes pooled together in analysis.
<sup>d</sup> HPV 16, 18, 31, 33, 35, 39, 45, 51, 52, 56, 58, 59, and 68.

**Figure 1.**
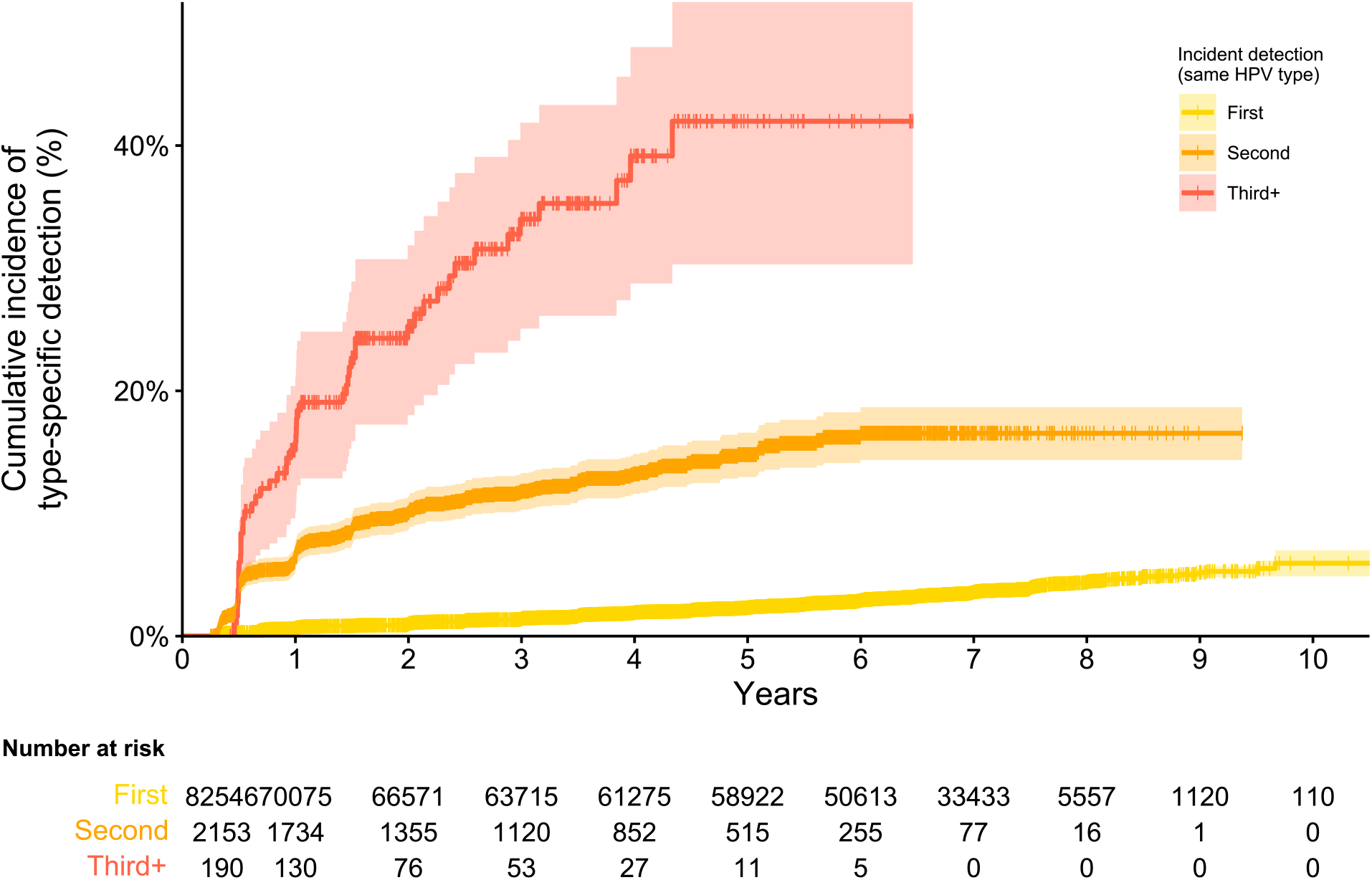
Cumulative incidence of genotype-specific incident detection of HPV (first incident detection) and of redetection after at least one negative visit of the same HPV genotype (second and third or more incident detection), pooled across all HPV genotypes. Time to detection is modeled from the baseline study visit for first HPV detection, and from the first negative visit following the prior detection of that genotype for redetections. Notches represent censored observations, and shaded regions represent 95% confidence intervals.

The joint test for the effect of HPV genotype suggested there were significant overall differences in incidence of first detection with different HPV genotypes (p<0.0001), with HPV16 being the genotype with the highest rate of first incident detection. There were also significant overall differences in the incidence of redetection with different HPV genotypes (p<0.0001), but while HPV16 had a higher redetection incidence than the pooled average (19% vs 15% at 5 years), there were many other genotypes with higher redetection incidences (Table 2).

While the rate of first incident detections decreased with age, age was not a significant predictor of the rate of re-detection (Table 3, Figure 2). The joint test for interaction was highly significant (p<0.0001), suggesting the effect of age is different for first incident detections than for re-detections. Menopausal status was not a significant predictor of either first incident detections or redetections. Being a current smoker was associated with a 1.32 (95%CI 0.99-1.76) times higher genotype-specific re-detection rate after adjustment for other variables. Increasing number of lifetime partners was associated with a significantly increased incidence of both first incident HPV detection and same genotype redetections. Having a sex partner in the last interval was not a significant predictor of either first incident detections or redetections; in most cases these were the woman’s ongoing steady sex partner. Conversely, having a new sex partner in the last interval was associated with a higher rate of first incident detections (HR 2.04, 95%CI 1.74-2.40), but not with same genotype redetections (HR 0.98, 95%CI 0.70-1.35); the joint test for interaction was significant (p<0.0001), suggesting the effect of having a new partner is different for first incident detections than for redetections.

**Table 3.** Hazard ratios of HPV genotype-specific first incident detection and re-detection (second or more detection) by women’s characteristics

| Variable<sup>a</sup> | First incident detection (N=2368) |  |  |  | Redetection (N=308) |  |  |  | Interaction<sup>c</sup><br>p-value |
| --- | --- | --- | --- | --- | --- | --- | --- | --- | --- |
|  | HR<br>(crude) | (95% CI) | HR<sup>b</sup><br>(adjusted) | (95% CI) | HR<br>(crude) | (95% CI) | HR<sup>b</sup><br>(adjusted) | (95% CI) |  |
| Age |  |  |  |  |  |  |  |  | <0.0001 |
| <25 | 1.00 | (ref) | 1.00 | (ref) | 1.00 | (ref) | 1.00 | (ref) |  |
| 25-34 | 0.53 | (0.44-0.62) | 0.57 | (0.49-0.67) | 0.81 | (0.56-1.18) | 0.82 | (0.56-1.19) |  |
| 35-44 | 0.34 | (0.28-0.41) | 0.38 | (0.31-0.46) | 1.20 | (0.82-1.77) | 1.18 | (0.79-1.77) |  |
| ≥45 | 0.32 | (0.25-0.40) | 0.36 | (0.28-0.45) | 0.87 | (0.54-1.39) | 0.90 | (0.54-1.47) |  |
| Menopausal status |  |  |  |  |  |  |  |  | 0.51 |
| Pre-menopausal | 1.00 | (ref) | 1.00 | (ref) | 1.00 | (ref) | 1.00 | (ref) |  |
| Post-menopausal | 0.89 | (0.66-1.19) | 1.30 | (0.96-1.76) | 0.98 | (0.44-2.19) | 0.98 | (0.44-2.22) |  |
| Income quartile |  |  |  |  |  |  |  |  | 0.48 |
| 1 (lowest) | 1.22 | (1.02-1.46) | 1.12 | (0.95-1.33) | 0.83 | (0.56-1.23) | 0.84 | (0.57-1.24) |  |
| 2 | 1.17 | (0.97-1.41) | 1.18 | (0.99-1.41) | 0.96 | (0.67-1.38) | 1.02 | (0.72-1.45) |  |
| 3 | 1.10 | (0.91-1.33) | 1.14 | (0.95-1.36) | 1.19 | (0.84-1.69) | 1.20 | (0.84-1.70) |  |
| 4 (highest) | 1.00 | (ref) | 1.00 | (ref) | 1.00 | (ref) | 1.00 | (ref) |  |
| Race |  |  |  |  |  |  |  |  | 0.42 |
| White | 1.00 | (ref) | 1.00 | (ref) | 1.00 | (ref) | 1.00 | (ref) |  |
| Non-white | 1.09 | (0.96-1.24) | 0.96 | (0.85-1.09) | 1.08 | (0.83-1.40) | 1.08 | (0.83-1.41) |  |
| Smoking status |  |  |  |  |  |  |  |  | 0.28 |
| Never | 1.00 | (ref) | 1.00 | (ref) | 1.00 | (ref) | 1.00 | (ref) |  |
| Former | 0.89 | (0.74-1.06) | 0.85 | (0.72-1.00) | 1.17 | (0.82-1.68) | 1.11 | (0.76-1.60) |  |
| Current | 1.11 | (0.97-1.28) | 1.06 | (0.93-1.22) | 1.44 | (1.09-1.91) | 1.32 | (0.99-1.76) |  |
| Condom use last interval |  |  |  |  |  |  |  |  | <0.0001 |
| Yes | 1.00 | (ref) | 1.00 | (ref) | 1.00 | (ref) | 1.00 | (ref) |  |
| No | 0.67 | (0.59-0.77) | 0.96 | (0.85-1.10) | 0.99 | (0.75-1.30) | 1.00 | (0.76-1.33) |  |
| Lifetime number of sex partners |  |  |  |  |  |  |  |  | <0.0001 |
| 0-1 | 1.00 | (ref) | 1.00 | (ref) | 1.00 | (ref) | 1.00 | (ref) |  |
| 2-3 | 1.83 | (1.58-2.12) | 1.65 | (1.42-1.91) | 1.17 | (0.82-1.67) | 1.16 | (0.82-1.65) |  |
| 4+ | 2.23 | (1.91-2.61) | 1.78 | (1.51-2.09) | 1.55 | (1.11-2.16) | 1.50 | (1.06-2.10) |  |
| Any sex partner in last interval |  |  |  |  |  |  |  |  | 0.88 |
| No | 1.00 | (ref) | 1.00 | (ref) | 1.00 | (ref) | 1.00 | (ref) |  |
| Yes | 1.06 | (0.89-1.27) | 0.89 | (0.73-1.08) | 0.94 | (0.61-1.45) | 0.92 | (0.59-1.45) |  |
| New sex partner in last interval |  |  |  |  |  |  |  |  | <0.0001 |
| No | 1.00 | (ref) | 1.00 | (ref) | 1.00 | (ref) | 1.00 | (ref) |  |
| Yes | 2.86 | (2.46-3.33) | 2.04 | (1.74-2.40) | 1.08 | (0.78-1.50) | 0.98 | (0.70-1.35) |  |

CI=confidence interval; HPV=human papillomavirus; HR=hazard ratio.
<sup>a</sup> All variables considered as time-varying predictors in analyses, except for income and race which were only measured at study baseline, and menopausal status which was only measured twice.
<sup>b</sup> Adjusted for all variables in table.
<sup>c</sup> Joint test for interaction effect between the predictor variable and order of detection (first or redetection) in the multivariable adjusted model using the robust sandwich variance estimate. This tests whether the HRs are equivalent for first incident detections and re-detections.

**Figure 2.**
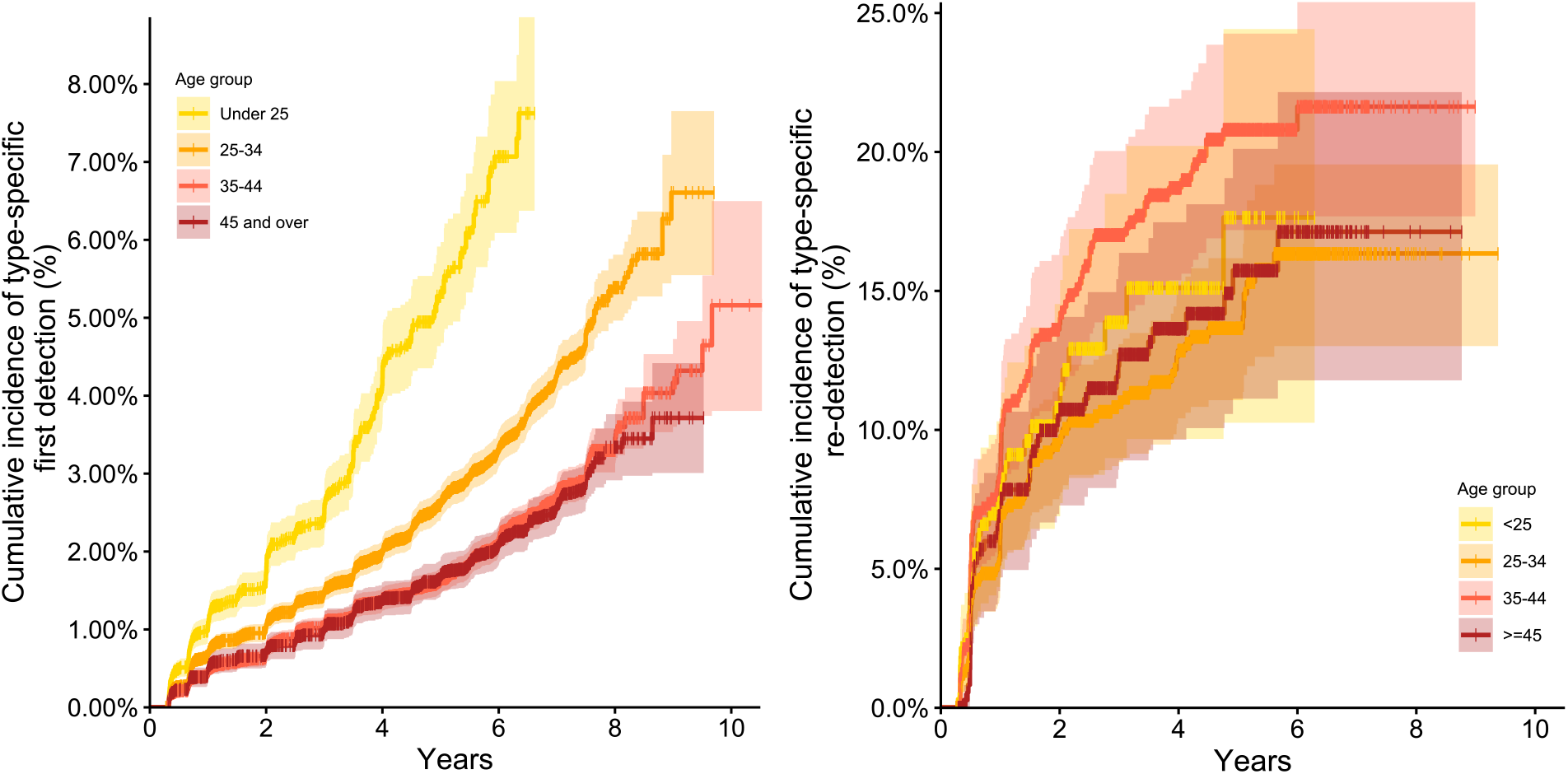
Cumulative incidence of HPV genotype-specific first detection (left) and redetection (right) by age at time of detection, pooled across all HPV genotypes. Time to detection is modeled from the baseline study visit for first HPV detection, and from the first negative visit following the prior detection of that genotype for redetection. Age is modeled as a time-varying exposure. Notches represent censored observations, and shaded regions represent 95% confidence intervals.

There were 646 ASCUS+ cytology results during follow-up, including 258 LSILs and 77 HSILs. Table 4 presents the prevalence of ASCUS+, LSIL, and HSIL across high-risk HPV genotype observations, i.e., the probability that a woman who is negative or positive for a given high-risk HPV genotype had a concurrent ASCUS+, LSIL, or HSIL cytology result. Because the unit of observation was the HPV genotype, numerators and denominators do not represent the number of cytological lesions over the number of women. Rather, they represent the number of genotype-specific observations across visits with prevalent cervical abnormalities over all genotype-specific observations across visits. The prevalence of HSIL was similar across visits with first detections (2.9%) and redetections (3.1%). Similar results were observed for LSIL and ASCUS+.

**Table 4.** Cross-sectional prevalence of cytological results by genotype-specific HPV positivity, pooled over all high-risk HPV genotypes and visits over all women.

| High-risk HPV status at visit (genotype-specific)<sup>a</sup> | ASCUS+ Prevalence |  | LSIL Prevalence |  | HSIL Prevalence |  |
| --- | --- | --- | --- | --- | --- | --- |
|  | n/Visits<sup>b</sup> | (%) | n/Visits<sup>b</sup> | (%) | n/Visits<sup>b</sup> | (%) |
| Negative | 7842/308004 | 2.6% | 3095/308004 | 1.0% | 928/308004 | 0.3% |
| Positive, first detection | 399/2264 | 17.6% | 202/2264 | 8.9% | 65/2264 | 2.9% |
| Positive, redetection | 40/250 | 16.0% | 18/250 | 7.2% | 8/250 | 3.2% |

ASCUS+= Atypical squamous cells of undetermined significance or higher; CI=confidence interval; HPV=human papillomavirus; HSIL=high grade squamous intraepithelial lesion; LSIL=low grade squamous intraepithelial lesion.
<sup>a</sup> High-risk HPV: 16, 18, 31, 33, 35, 39, 45, 51, 52, 56, 58, 59, 66, and 68.
<sup>b</sup> Numerator is number of genotype-specific visits with given cytological results over all women, denominator is number of genotype-specific visits over all women.

## Discussion

In this analysis of the Ludwig-McGill cohort over a median follow-up of 6.5 years, many women had intermittent detection patterns with individual HPV genotypes. The risk of redetection of a given HPV genotype was 15% by 5 years after the date a woman became negative for that genotype, pooled across HPV genotypes. While first HPV detections were associated with the acquisition of new sex partners, same genotype HPV redetections were not. We observed a similar prevalence of HSIL at visits with first detections and redetections with the same HPV genotype.

Our results suggest that many HPV redetections are likely to be reactivations of latent infections as well as new reinfections from sexual exposure. Previous studies have found that many HPV infections can have intermittent detection patterns.[19-23] These tend to have a lower viral load than persistent or transient detections,[8,21] suggesting that some of these redetections may be infections with viral shedding fluctuating below the limit of detection. We found that 9% of genotype-specific HPV-positive observations in the Ludwig-McGill cohort study had intermittent detection patterns. This was similar to a cohort study from Costa Rica which found that 8% of women ever HPV positive over 7 years had intermittent detection patterns,[20] but lower than studies from the USA which found that 13-26% of HPV detections had intermittent detection patterns.[21,24] However, the above results are based on crude counts rather than survival analyses; the proportion of observations with intermittent detection patterns may vary across studies with different follow-up due to right-censoring of observations. Survival analysis methods provide a better estimate of the cumulative probability of redetection over time by accounting for censoring. Using a Cox model, we estimated that 5 years after loss of positivity of a first HPV detection there was a 15% cumulative probability of redetection of the same HPV genotype. These results are similar to another study in young US women which found that 18% of HPV16 infections became redetectable by 8.5 years after the date of loss of positivity.[25] Redetection of the same HPV genotype is therefore fairly common and can occur several years after testing negative.

While having new sexual partners was associated with an increased risk of first HPV detection, it was not associated with redetections of the same HPV genotype in our analysis. The effect of new sexual partner acquisition varies across studies, with some finding that having new sexual partners increases the risk of redetection[8,25] while others do not.[23] Our results should be interpreted in light of the previous analysis by Trottier *et al*. of the Ludwig-McGill cohort study,[8] which had found that some redetections were reinfections associated with the acquisition of new sexual partners. There were fundamental differences between analyses that should be noted. The Trottier *et al*. study had research questions focused on HPV reinfections, and restricted analyses to redetections occurring after three consecutive negative visits in order to exclude potential intermittent detection. In contrast, we included all redetections with at least one intervening negative result; only 30% (92/308) of genotype-specific redetections in our analysis occurred after 3 intervening negative results (Supplementary Table 1). This restriction accounts for part of the difference in results. The other major difference was that Trottier *et al*. performed a woman-level analysis rather than an HPV genotype-level analysis. Woman-level analyses are closer to the results that a woman would receive in the clinic using a screening test targeting multiple HPV genotypes at once, whereas HPV genotype-level analyses focus on understanding the biological course of individual infections. Trottier *et al*.’s woman-level analysis looked at the time to first redetection with any HPV genotype a woman was previously positive for, so was restricted to 19 first redetections and excluded 73 other same-genotype redetections after 3 negative intervening results. These first redetections across multiple genotypes were more strongly associated with new sexual partner acquisition, suggesting these are a subsample more likely to be true new reinfections from sexual transmission. This effect of new sexual partners on HPV redetection was strongly diluted in our current analysis due to the inclusion of many more genotype-specific redetections than in Trottier *et al*. The combined results from both analyses suggest that while there are redetections that are true new infections caused by sexual transmission, the majority of genotype-specific HPV redetections in this cohort are not associated with new partner acquisition, and may instead represent reactivations/redetections of previously existing infections.

It is unclear whether HPV redetections are associated with the same risk of cytological lesions as first detections. The Guanacaste cohort study had found that women with HPV detected only once and women with HPV redetections both had a similar 7-9% risk of CIN2+.[20] Results from Kaiser Permanente Northern California found that the risk of CIN3+ in women with intermittent detection patterns depended mostly on whether the woman had a current HPV positive result.[4] We found that the prevalence of cytological lesions did not substantially differ between first and subsequent redetection episodes with the same HPV genotype. However, there were very few HSILs detected in this population, so the statistical power was low for assessing the risk of HSIL.

Increasing age was associated with a lower risk of first HPV detection but not with HPV redetections. This was expected for first detections, as age is correlated with sexual activity and may also be a proxy of the prevalence of HPV infection in women’s sexual partners. The lack of association between age and redetections conversely suggests that redetection is less likely to be attributable to sexual transmission. These results are also not consistent with the hypothesis that the risk of reactivation of latent infections increases with age. While increasing age is associated with a decline in immune responses to pathogens,[26] the age range of participants in the Ludwig-McGill cohort study may have been too narrow to detect any potential effects of immunosenescence on HPV reactivation.

A potential limitation of our study was that because women were recruited from a maternal and child health program, participants are likely to be a sample at lower risk of new HPV infections than the general population. The fraction of all HPV detections attributable to sexual transmission may be lower in this population, as most women were married or living as married with a steady partner, and few reported new sexual partners over the course of the study. The fraction of detections attributable to sexual transmission is likely to be higher in populations with more sexual partner turnover.[27,28]

In summary, we found that redetection of the same HPV genotype is fairly common in women and may in some cases occur many years after a woman becomes HPV DNA negative. These results may help in de-stigmatizing a positive HPV test result, as they suggest that many HPV detections may be a reactivated past infection, rather than a new infection from recent sexual behaviors or partner infidelity. The results also suggest that episodes of HPV redetection are associated with a similar risk of underlying cervical lesions as first detections.

## Data Availability

Participants of the Ludwig-McGill cohort study did not consent to have their data made publicly available, and confidentiality precludes the publishing of their data. To access the data for research purposes, please contact Eduardo Franco or Luisa Villa. The program code for the current results is available at the McGill University Dataverse repository: https://doi.org/10.5683/SP3/EIF6A7.

## Acknowledgements

Ludwig-McGill Cohort Study Team Members: Affiliated with the Ludwig Institute for Cancer Research in Sao Paulo, Brazil: Maria Luiza Baggio, Lenice Galan, João Simão Sobrinho, José Carlos Mann Prado, Lara Termini, Maria Cecília Costa, Romulo Miyamura, Andrea Trevisan, Patricia Thomann, João Candeias, Laura Sichero, Paula Rahal, Antonio Ruiz, Jane Kaiano, Monica Santos, Patricia Savio, Paulo Maciag, Tatiana Rabachini, Silvaneide Ferreira, Luisa Villa (co-principal investigator). Affiliated with McGill University in Montreal, Canada: Mariam El-Zein, Marie-Claude Rousseau, Salaheddin Mahmud, Nicolas Schlecht, Helen Trottier, Harriet Richardson, Alex Ferenczy, Thomas Rohan, Myriam Chevarie-Davis, Karolina Louvanto, Joseph Tota, Eileen Shaw, Agnihotram Ramanakumar, Eliane Duarte, Sophie Kulaga, Juliette Robitaille, Eduardo Franco (principal investigator).

