## Supplementary Appendix for "Human papillomavirus intermittence, redetections, and associated risk of cytological abnormalities in the Ludwig-McGill cohort study of adult women"

Supplementary Table 1. HPV genotype-specific detection patterns in women with at least 3 study visits (1=positive visit, 0=negative visit).

| Pattern | Times the pattern was observed | Pattern | Times the pattern was observed | Pattern | Times the pattern was observed |
| --- | --- | --- | --- | --- | --- |
| Prevalent and persistent |  | **Prevalent and transient** |  | **Prevalent and intermittent** |  |
| 110 | 4 | 100 | 11 | 10000000000010 | 1 |
| 1100 | 3 | 1000 | 7 | 1000000000010 | 1 |
| 1100000 | 1 | 10000 | 2 | 10000000001000 | 1 |
| 110000000 | 3 | 100000 | 5 | 100000010000 | 1 |
| 1100000000 | 5 | 1000000 | 7 | 10000001000000 | 1 |
| 11000000000 | 4 | 10000000 | 4 | 100000100000 | 1 |
| 110000000000 | 10 | 100000000 | 3 | 1000001000000 | 1 |
| 1100000000000 | 14 | 1000000000 | 12 | 100000110000 | 1 |
| 11000000000000 | 13 | 10000000000 | 8 | 1000010000000 | 1 |
| 110000000000000 | 7 | 100000000000 | 26 | 10001111 | 1 |
| 1100000000000000 | 1 | 1000000000000 | 36 | 10010000000000 | 1 |
| 111 | 4 | 10000000000000 | 65 | 10010100 | 1 |
| 11100 | 1 | 100000000000000 | 17 | 100101111111 | 1 |
| 111000 | 1 | 1000000000000000 | 2 | 10011100000 | 1 |
| 1110000 | 1 |  |  | 101 | 2 |
| 111000000 | 1 |  |  | 10100000000000 | 3 |
| 1110000000 | 3 |  |  | 101000000000000 | 3 |
| 111000000000 | 2 |  |  | 1010001110 | 1 |
| 1110000000000 | 7 |  |  | 10101 | 1 |
| 11100000000000 | 11 |  |  | 10101100 | 1 |
| 111000000000000 | 2 |  |  | 10110 | 1 |
| 1111 | 7 |  |  | 1011000000000 | 1 |
| 111100000 | 1 |  |  | 10110010000000 | 1 |
| 11110000000 | 1 |  |  | 10111000000000 | 3 |
| 111100000000 | 2 |  |  | 10111010 | 1 |
| 1111000000000 | 3 |  |  | 10111110000000 | 1 |
| 11110000000000 | 7 |  |  | 1100000000001 | 1 |
| 111110 | 1 |  |  | 1100000000100 | 1 |
| 11111000000000 | 2 |  |  | 1100000010100 | 1 |
| 111111 | 1 |  |  | 11000100000000 | 1 |
| 11111100000 | 1 |  |  | 110001110000000 | 1 |
| 111111000000000 | 1 |  |  | 1100101000000 | 1 |
| 111111100000 | 1 |  |  | 11001011111111 | 1 |
| 1111111000000 | 1 |  |  | 1101000000000 | 1 |
| 111111100000000 | 1 |  |  | 11010000000000 | 2 |
| 11111111 | 2 |  |  | 1101000000010 | 1 |
| 1111111100000 | 1 |  |  | 110100100 | 1 |
| 111111111 | 2 |  |  | 11011 | 1 |
| 11111111111000 | 1 |  |  | 1101100000000 | 1 |
| 111111111111 | 2 |  |  | 1101111000000 | 1 |
| 1111111111111 | 1 |  |  | 1110001000001 | 1 |
|  |  |  |  | 1110011100001 | 1 |
|  |  |  |  | 111010000000 | 1 |
|  |  |  |  | 11101100000110 | 1 |
|  |  |  |  | 111100000001 | 1 |
|  |  |  |  | 1111011 | 1 |
|  |  |  |  | 11110111100111 | 1 |
|  |  |  |  | 11111001100000 | 1 |
|  |  |  |  | 1111101000000 | 1 |
|  |  |  |  | 1111101110000 | 1 |
|  |  |  |  | 111110111010110 | 1 |
|  |  |  |  | 1111111001110 | 1 |
| Incident and persistent |  | **Incident and transient** |  | **Incident and intermittent** |  |
| 000000000000011 | 8 | 000000000000001 | 18 | 000000000001001 | 1 |
| 00000000000011 | 18 | 0000000000000010 | 3 | 00000000000101 | 6 |
| 000000000000110 | 2 | 00000000000001 | 79 | 000000000010001 | 1 |
| 000000000000111 | 5 | 000000000000010 | 18 | 00000000001001 | 4 |
| 0000000000011 | 9 | 0000000000001 | 30 | 0000000000101 | 2 |
| 00000000000110 | 17 | 00000000000010 | 55 | 00000000001010 | 1 |
| 000000000001100 | 2 | 000000000000100 | 23 | 00000000001011 | 4 |
| 00000000000111 | 6 | 000000000001 | 25 | 000000000010110 | 1 |
| 000000000001110 | 3 | 0000000000010 | 23 | 00000000001101 | 1 |
| 000000000011 | 10 | 00000000000100 | 51 | 000000000100010 | 1 |
| 0000000000110 | 6 | 000000000001000 | 20 | 0000000001001 | 1 |
| 00000000001100 | 10 | 0000000000010000 | 1 | 00000000010010 | 1 |
| 000000000011000 | 3 | 00000000001 | 9 | 000000000100100 | 1 |
| 0000000000111 | 5 | 000000000010 | 11 | 00000000010100 | 4 |
| 00000000001110 | 5 | 0000000000100 | 32 | 000000000101000 | 1 |
| 000000000011100 | 2 | 00000000001000 | 94 | 00000000011001 | 1 |
| 00000000001111 | 8 | 000000000010000 | 20 | 00000000011011 | 2 |
| 000000000011110 | 2 | 0000000001 | 3 | 000000001000001 | 1 |
| 000000000011111 | 1 | 00000000010 | 10 | 000000001000010 | 1 |
| 00000000011 | 2 | 000000000100 | 8 | 000000001001 | 1 |
| 000000000110 | 3 | 0000000001000 | 20 | 00000000100100 | 1 |
| 0000000001100 | 4 | 00000000010000 | 42 | 00000000100101 | 1 |
| 00000000011000 | 6 | 000000000100000 | 14 | 00000000100110 | 1 |
| 000000000110000 | 3 | 000000001 | 3 | 00000000101 | 1 |
| 000000000111 | 3 | 0000000010 | 3 | 0000000010100 | 1 |
| 0000000001110 | 1 | 00000000100 | 11 | 00000000101000 | 1 |
| 00000000011100 | 2 | 000000001000 | 12 | 000000001010000 | 2 |
| 000000000111000 | 2 | 0000000010000 | 13 | 00000000101011 | 1 |
| 0000000001111 | 1 | 00000000100000 | 38 | 000000001011001 | 1 |
| 00000000011110 | 1 | 000000001000000 | 14 | 000000001110101 | 1 |
| 00000000011111 | 4 | 00000001 | 14 | 0000000100011 | 1 |
| 000000000111111 | 1 | 000000010 | 8 | 00000001000110 | 1 |
| 0000000011 | 5 | 0000000100 | 5 | 0000000100100 | 1 |
| 0000000011000 | 4 | 00000001000 | 6 | 00000001001000 | 1 |
| 00000000110000 | 15 | 000000010000 | 16 | 000000010010000 | 1 |
| 000000001100000 | 5 | 0000000100000 | 27 | 000000011001 | 1 |
| 000000001110 | 1 | 00000001000000 | 37 | 000000011001111 | 1 |
| 0000000011100 | 2 | 000000010000000 | 20 | 0000000111010 | 1 |
| 00000000111000 | 6 | 0000000100000000 | 2 | 00000001110110 | 1 |
| 000000001110000 | 1 | 0000001 | 9 | 00000001111101 | 1 |
| 00000000111100 | 1 | 00000010 | 6 | 000000100000001 | 1 |
| 0000000011111 | 1 | 000000100 | 4 | 00000010000001 | 1 |
| 000000001111100 | 1 | 0000001000 | 6 | 0000001000001 | 1 |
| 00000000111111 | 2 | 00000010000 | 6 | 000000100000100 | 1 |
| 000000011 | 5 | 000000100000 | 15 | 00000010000100 | 1 |
| 0000000110 | 1 | 0000001000000 | 19 | 0000001000100 | 2 |
| 00000001100 | 3 | 00000010000000 | 53 | 000000100100 | 1 |
| 000000011000 | 5 | 000000100000000 | 10 | 00000010011100 | 1 |
| 0000000110000 | 3 | 000001 | 1 | 0000001010000 | 2 |
| 00000001100000 | 13 | 0000010 | 4 | 00000010100000 | 2 |
| 000000011000000 | 1 | 00000100 | 1 | 00000011010 | 1 |
| 0000000111 | 1 | 000001000 | 5 | 000000110100 | 1 |
| 000000011100 | 3 | 0000010000 | 4 | 00000011010000 | 1 |
| 0000000111000 | 1 | 00000100000 | 5 | 00000011100100 | 1 |
| 00000001110000 | 2 | 000001000000 | 11 | 000000111010 | 1 |
| 000000011111 | 1 | 0000010000000 | 16 | 000000111010000 | 1 |
| 0000000111110 | 2 | 00000100000000 | 36 | 00000011110100 | 1 |
| 00000001111111 | 3 | 000001000000000 | 17 | 00000011110111 | 1 |
| 000000110 | 1 | 00001 | 4 | 00000100000010 | 2 |
| 0000001100 | 1 | 000010 | 2 | 00000100001000 | 1 |
| 000000110000 | 1 | 0000100 | 9 | 000001000010000 | 1 |
| 0000001100000 | 1 | 00001000 | 4 | 00000100010000 | 1 |
| 00000011000000 | 12 | 000010000 | 6 | 000001001 | 1 |
| 000000110000000 | 1 | 0000100000 | 4 | 00000100100100 | 1 |
| 0000001100000000 | 1 | 00001000000 | 5 | 00000100100110 | 1 |
| 000000111 | 1 | 000010000000 | 17 | 0000010011 | 1 |
| 0000001110 | 1 | 0000100000000 | 22 | 000001001111 | 1 |
| 00000011100000 | 2 | 00001000000000 | 71 | 0000010100000 | 1 |
| 000000111000000 | 2 | 000010000000000 | 20 | 00000101000000 | 1 |
| 0000001111 | 2 | 0000100000000000 | 1 | 000001010000000 | 1 |
| 00000011111 | 1 | 0001 | 8 | 000001011 | 1 |
| 000000111111 | 1 | 000100 | 3 | 00000101100000 | 2 |
| 0000001111111 | 1 | 0001000 | 3 | 0000011000010 | 1 |
| 00000011111111 | 1 | 00010000 | 4 | 00000110110101 | 1 |
| 0000001111111111 | 1 | 000100000 | 2 | 00000111010000 | 2 |
| 0000011 | 2 | 0001000000 | 8 | 000001111000001 | 1 |
| 000001100 | 1 | 00010000000 | 10 | 00000111110001 | 1 |
| 0000011000 | 1 | 000100000000 | 18 | 00001000000001 | 1 |
| 00000110000 | 1 | 0001000000000 | 21 | 00001000000010 | 1 |
| 000001100000 | 4 | 00010000000000 | 39 | 00001000001111 | 1 |
| 00000110000000 | 13 | 000100000000000 | 8 | 0000100001 | 1 |
| 000001100000000 | 4 | 0001000000000000 | 2 | 000010001 | 1 |
| 0000011100 | 1 | 001 | 19 | 0000100010 | 1 |
| 000001110000 | 2 | 0010 | 6 | 00001000100000 | 1 |
| 0000011100000 | 2 | 00100 | 2 | 00001000110000 | 1 |
| 00000111000000 | 4 | 001000 | 2 | 000010001101000 | 1 |
| 000001110000000 | 3 | 0010000 | 2 | 000010010 | 1 |
| 00000111100000 | 1 | 00100000 | 5 | 00001001000000 | 1 |
| 0000011110000000 | 1 | 001000000 | 8 | 000010010000000 | 1 |
| 000001111100000 | 1 | 0010000000 | 5 | 0000100111010 | 1 |
| 000001111110 | 1 | 00100000000 | 4 | 00001001111111 | 1 |
| 00000111111000 | 1 | 001000000000 | 20 | 00001010000000 | 2 |
| 000011 | 1 | 0010000000000 | 31 | 000010100100000 | 1 |
| 0000110 | 1 | 00100000000000 | 38 | 00001011000 | 1 |
| 0000110000 | 1 | 001000000000000 | 9 | 00001100000001 | 1 |
| 00001100000 | 1 | 010 | 14 | 000011000100 | 1 |
| 000011000000 | 5 | 0100 | 8 | 00001101100001 | 1 |
| 0000110000000 | 2 | 01000 | 3 | 000100000000100 | 1 |
| 00001100000000 | 8 | 010000 | 4 | 00010000000100 | 1 |
| 000011000000000 | 3 | 0100000 | 4 | 000100000001000 | 1 |
| 0000111 | 1 | 01000000 | 2 | 000100000001101 | 1 |
| 000011100 | 1 | 010000000 | 3 | 00010000001010 | 1 |
| 0000111000 | 1 | 0100000000 | 10 | 000100000110 | 1 |
| 000011100000 | 4 | 01000000000 | 6 | 000100001001 | 1 |
| 0000111100 | 1 | 010000000000 | 13 | 000100001001000 | 1 |
| 000011110000 | 1 | 0100000000000 | 23 | 000100011000001 | 1 |
| 0000111100000 | 1 | 01000000000000 | 45 | 00010001101001 | 1 |
| 00001111000000 | 2 | 010000000000000 | 11 | 00010001110001 | 1 |
| 000011110000000 | 1 | 0100000000000000 | 2 | 00010010000000 | 1 |
| 00001111100 | 1 |  |  | 00010011100000 | 1 |
| 0000111110000 | 1 |  |  | 00010100000 | 1 |
| 00001111100000 | 1 |  |  | 0001010000000 | 3 |
| 00001111110000 | 1 |  |  | 00010100000111 | 1 |
| 000011111111 | 1 |  |  | 00010101110000 | 1 |
| 0000111111111 | 1 |  |  | 000101111000000 | 1 |
| 00001111111111 | 1 |  |  | 000110100 | 1 |
| 000011111111111 | 1 |  |  | 00011110011 | 1 |
| 000110 | 1 |  |  | 00100000000010 | 2 |
| 000110000 | 1 |  |  | 00100000000100 | 1 |
| 0001100000 | 1 |  |  | 00100000010000 | 1 |
| 000110000000 | 1 |  |  | 001000001000 | 1 |
| 0001100000000 | 2 |  |  | 00100001000000 | 1 |
| 00011000000000 | 5 |  |  | 001000100 | 1 |
| 000111000000 | 1 |  |  | 0010001000000 | 1 |
| 0001110000000 | 1 |  |  | 001001000000000 | 1 |
| 0001111000000 | 2 |  |  | 00100100011111 | 1 |
| 00011110000000 | 2 |  |  | 00100111000010 | 1 |
| 000111100000000 | 1 |  |  | 0010011110000 | 1 |
| 00011111 | 1 |  |  | 001010000000 | 1 |
| 000111110000000 | 1 |  |  | 0010100000000 | 1 |
| 00011111100000 | 1 |  |  | 00101000000000 | 1 |
| 0011 | 2 |  |  | 00101011111111 | 1 |
| 00110 | 1 |  |  | 0010110000 | 1 |
| 001100 | 1 |  |  | 00101100000000 | 1 |
| 0011000 | 3 |  |  | 00101110000010 | 1 |
| 001100000 | 1 |  |  | 001011111100111 | 1 |
| 0011000000 | 2 |  |  | 00110001000000 | 1 |
| 00110000000 | 1 |  |  | 00110100000 | 1 |
| 001100000000 | 3 |  |  | 001101111 | 1 |
| 0011000000000 | 7 |  |  | 001111001 | 1 |
| 00110000000000 | 13 |  |  | 0011110111 | 1 |
| 001100000000000 | 5 |  |  | 01000000010000 | 2 |
| 00111 | 1 |  |  | 010000001100 | 1 |
| 0011100000000 | 1 |  |  | 010000011111 | 1 |
| 00111000000000 | 2 |  |  | 01000010000 | 1 |
| 001110000000000 | 1 |  |  | 01000011010000 | 1 |
| 00111100000 | 1 |  |  | 01000110000 | 1 |
| 001111000000 | 1 |  |  | 01010000 | 2 |
| 001111100000000 | 1 |  |  | 0101000000000 | 2 |
| 00111111000000 | 1 |  |  | 01010000000000 | 3 |
| 0011111111 | 1 |  |  | 010100000100 | 1 |
| 0011111111111 | 1 |  |  | 010110 | 1 |
| 00111111111110 | 1 |  |  | 0101101000100 | 1 |
| 001111111111111 | 1 |  |  | 01011100000000 | 1 |
| 011 | 4 |  |  | 011000000100 | 1 |
| 0110 | 1 |  |  | 01100100000000 | 1 |
| 01100 | 1 |  |  | 01101011 | 1 |
| 0110000 | 1 |  |  | 01101111111110 | 1 |
| 0110000000 | 1 |  |  | 01110010000000 | 1 |
| 01100000000 | 4 |  |  | 01110100000 | 1 |
| 011000000000 | 4 |  |  | 011101000000 | 1 |
| 0110000000000 | 4 |  |  | 01111000000010 | 1 |
| 01100000000000 | 9 |  |  | 01111110011000 | 1 |
| 011000000000000 | 2 |  |  | 011111101110101 | 1 |
| 0111 | 4 |  |  |  |  |
| 011100 | 1 |  |  |  |  |
| 0111000 | 1 |  |  |  |  |
| 0111000000 | 2 |  |  |  |  |
| 01110000000 | 1 |  |  |  |  |
| 011100000000 | 2 |  |  |  |  |
| 0111000000000 | 1 |  |  |  |  |
| 01110000000000 | 5 |  |  |  |  |
| 011100000000000 | 1 |  |  |  |  |
| 011110 | 1 |  |  |  |  |
| 0111100 | 1 |  |  |  |  |
| 01111000000 | 1 |  |  |  |  |
| 011110000000 | 1 |  |  |  |  |
| 01111000000000 | 1 |  |  |  |  |
| 011110000000000 | 2 |  |  |  |  |
| 0111110 | 1 |  |  |  |  |
| 011111000000 | 1 |  |  |  |  |
| 0111111 | 1 |  |  |  |  |
| 01111110 | 1 |  |  |  |  |
| 01111110000000 | 1 |  |  |  |  |
